# Cannabidivarin for the treatment of HIV-associated neuropathic pain – a randomized, blinded, controlled clinical trial

**DOI:** 10.1101/2019.12.20.19015495

**Authors:** Luca Eibach, Simone Scheffel, Madeleine Cardebring, Marie Lettau, Özgür Celik, Andreas Morguet, Robert Röhle, Christoph Stein

## Abstract

**Background:** HIV remains a major burden to the health care system and neuropathic pain is the most common neurological complication of HIV-infection. Since current treatment strategies often lack satisfying pain relief, cannabinoids are discussed as a new option. We investigated Cannabidivarin as treatment for HIV-associated neuropathic pain.

**Methods:** We conducted a randomized, double-blind, placebo-controlled cross-over study. Patients underwent two successive treatment phases (4 weeks each) and were treated with Cannabidivarin (400mg/d) or placebo in a randomized order. A 3-week wash-out phase was designed to eliminate potential carry-overeffects and patients were followed up for 3 weeks after the end of the second treatment phase. The primary endpoint was pain intensity on an 11-point numeric rating scale and was recorded in a diary. Secondary endpoints were additional pain medication, pain characteristics and quality of life.

**Results:** We included 32 (31 male) patients. The mean pain intensity under Cannabidivarin was by 0.62 points higher compared to placebo (p=0.16; 95% CI -0.27 to 1.51). Cannabidivarin did not influence the amount of additional pain medication, pain characteristics or quality of life. No suspected unexpected adverse reactions occurred during the trial.

**Discussion:** Cannabidivarin was safe but failed to reduce neuropathic pain intensity in HIV-patients. This may be explained by a lack of cannabinoid receptor activation, as indicated by preclinical experiments. Further studies on larger sample sizes are needed.

## Introduction

Approximately 7-8% of the general population suffer from neuropathic pain, defined as ’pain that arises as a direct consequence of lesions or diseases affecting the somatosensory system’(1,2). Chronic neuropathic pain impairs quality of life and negatively affects the patients’ social relationships (3). Among various diseases that can underlie neuropathic pain, HIV-infection belongs to the most prevalent (4) and despite the development of highly effective antiretroviral therapy, HIV remains a major burden to the health system (5).

HIV-associated neuropathic pain usually occurs together with distal sensory neuropathy with symptoms of burning or dysesthesia in combination with numbness in stocking- or glove-like distribution (6), and may be caused by the inflammatory effects of HIV-infected macrophages and other neurodegenerative mechanisms (6,7). Furthermore, antiretroviral drugs, mainly dideoxynucleoside reverse transcriptase inhibitors, can cause mitochondrial and nerve damage (7,8) so that they are no longer recommended (9). Despite novel, more effective and less neurotoxic antiretroviral drugs, the prevalence of neuropathic pain in HIV-infected patients is still high and causal treatment is not available (6). Although treatment of chronic neuropathic pain should be based on both pharmacological and interdisciplinary non-pharmacological approaches (e.g. behavioral, physical and/or occupational therapy) (4), pharmacological therapy often predominates. Antidepressants, anticonvulsants and opioid analgesics are medications of choice (10). However, they often lack efficacy (4) and are limited by side effects such as respiratory depression, addiction and sedative effects (11), resulting in extensive additional costs and reduced quality of life (3,12).

Endocannabinoids, e.g. 2-arachidonylglycerol and anandamide, influence the transmission of pain signals by acting on cannabinoid (CB)-receptors 1 and 2 (13,14). Some exogenous cannabinoids have shown promising results in the treatment of neuropathic pain but they were limited by complicated dosing of smoked cannabis and side effects like nausea or drowsiness (15,16). Therefore, improved cannabinoid and opioid analgesics are being developed (10,13,14,17).

In this study, we investigated Cannabidivarin (CBDV) a novel phytocannabinoid derived from the *Cannabis sativa* L. plant, in patients with HIV-associated neuropathic pain. Using a double-blind cross-over trial design, we assessed pain, side effects and quality of life, and sought to correlate treatment responses to the patients’ genotype.

## Results

### Patient population

From January 2015 to September 2018 a total of 194 patients were contacted by email or phone, of which 55 were screened in the study center. 34 patients were initially enrolled and assigned a patient ID. The data of two patients could not be used for final efficacy analysis due to missing data or screening failure but were included in the safety population (**Figure 1**). Characteristics of the remaining 32 patients are shown in **Table 1**. All patients met the inclusion criterion of a positive Douleur Neuropathique 4 interview (DN4i) (≥3). Of the remaining 32 patients, 4 dropped-out during the study but were not excluded from analysis. The data of 28 patients (27 male, 1 female) were used for full efficacy analysis. Patients were randomized to receive CBDV in treatment phase A followed by placebo in treatment phase B (C-P), or placebo in phase A followed by CBDV in phase B (P-C).

**Table 1.** Data at day of initial screening. Abbreviations: C-P = CBDV-placebo; P-C = placebo-CBDV; y = years; cART = combined Antiretroviral Therapy, SD = standard deviation; NRS = Numeric Rating Scale

| Sequence of treatments |  | C-P | P-C | Total |
| --- | --- | --- | --- | --- |
| Male, n (%) |  | 16 (100) | 15 (93.8) | 31 (96.9) |
| Female, n |  | 0 | 1 (6.2) | 1 (3.1) |
| Age, y | mean (SD) | 52.31 (8.06) | 48.31 (9.62) | 50.31 (8.96) |
|  | range | 36 - 65 | 31 - 65 | 31 - 65 |
| NRS score, (0-10) | mean (SD) | 6.12 (1.15) | 6.44 (1.59) | 6.28 (1.37) |
|  | range | 4 - 8 | 4 - 9 | 4 - 9 |
| DN4i, (0-7) | mean (SD) | 5.19 (1.17) | 5 (0.89) | 5.09 (1.03) |
|  | range | 3 - 7 | 4 - 6 | 3 - 7 |
| Duration of pain, y | mean (SD) | 16.47 (7.91) | 9.94 (8.77) | 13.1 (8.87) |
|  | range | 2 - 30 | 1 - 27 | 1 - 30 |
| Duration of HIV infection, y | mean (SD) | 24.88 (9.17) | 17.81 (10.81) | 21.4 (10.2) |
|  | range | 3 - 33 | 2 - 32 | 2 - 33 |
| On cART, n |  | 16 | 15 | 31 |

**Figure 1.**
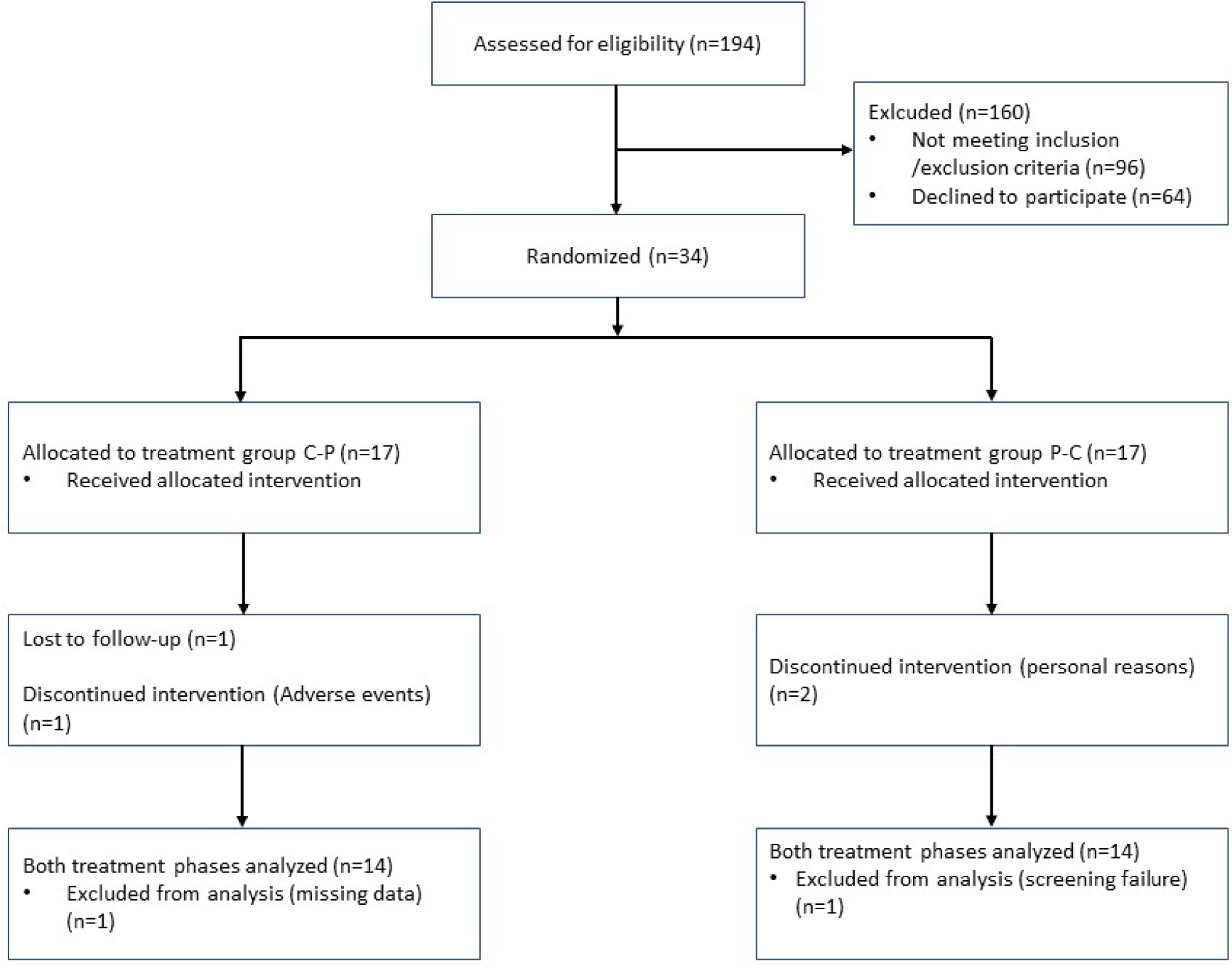
Flow chart.

### Primary endpoint

Overall, mean pain intensity (NRS; numeric rating scale) at the end of CBDV treatment was 0.62 points higher compared to placebo; this difference was not significant (p=0.16; 95% CI -0.27 to 1.51) (**Figure 2, Figure 3 and Table S 1**) The mean NRS value at the end of follow-up (3 weeks after end of treatment phase B) was 2.74 (SD: 1.47) in the C-P group and 3.67 (SD: 2.62) in the P-C group. During CBDV treatment, 9 patients experienced a mean pain reduction of at least 20 % and were therefore classified as CBDV responders. By the same criteria, 19 patients were classified as placebo responders.

**Figure 2.**
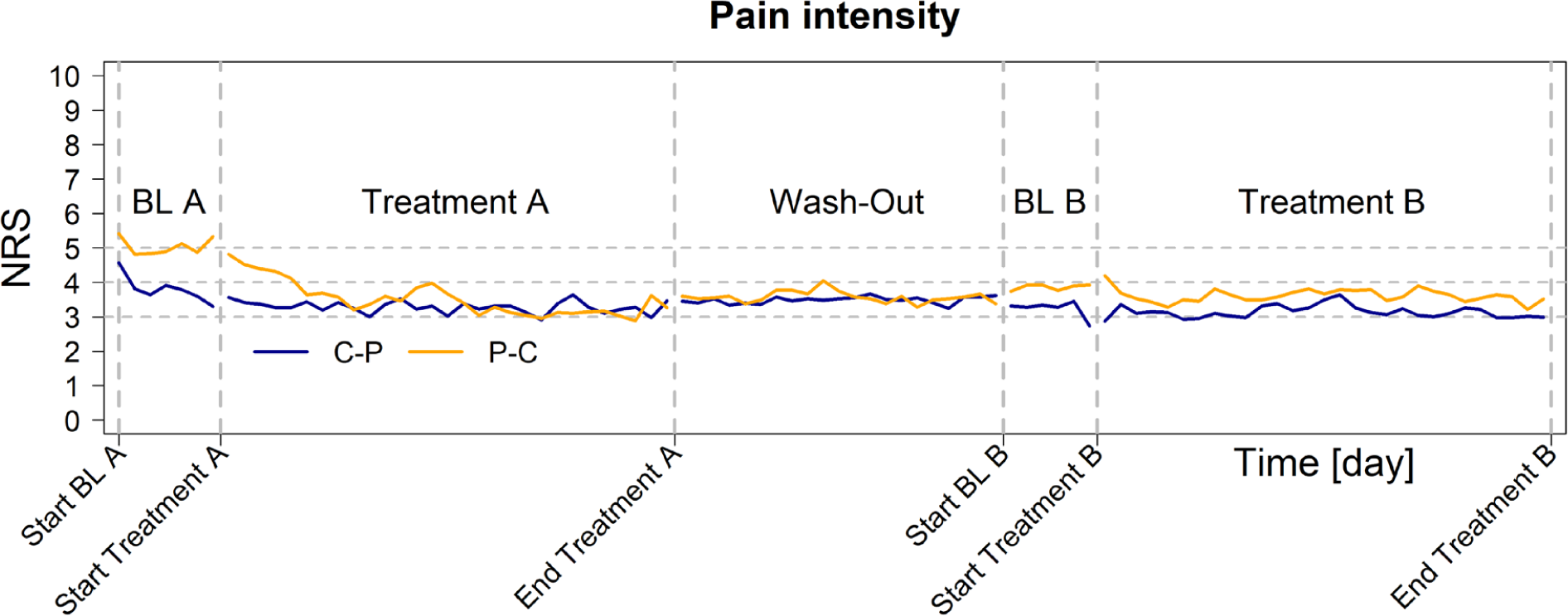
Pain intensity over time: Mean intensities per day for each group by treatment sequence. Abbreviations: C-P = CBDV-placebo; P-C = placebo-CBDV; BL= Baseline NRS = Numeric Rating Scale

**Figure 3.**
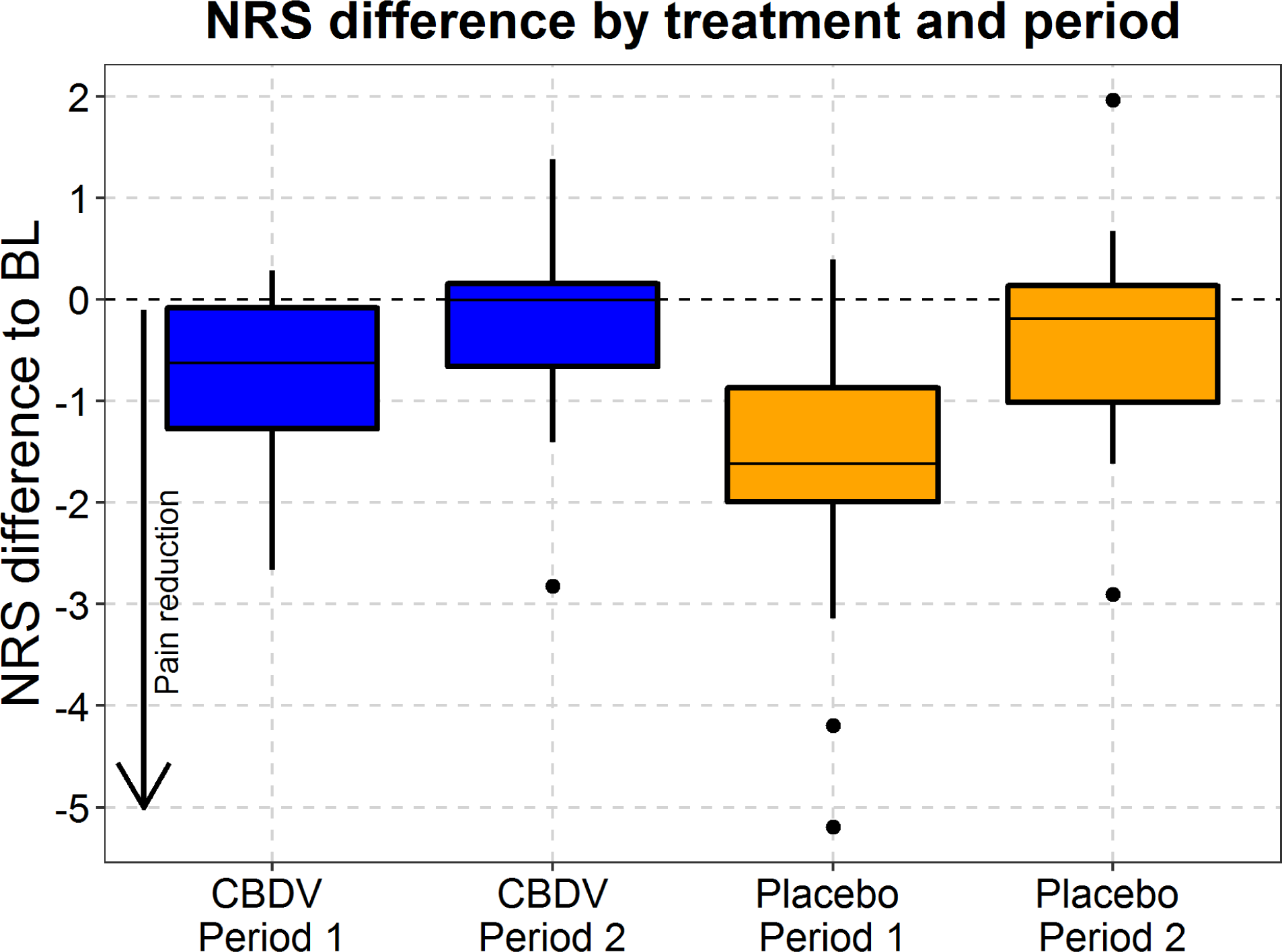
Boxplots of NRS values under CBDV and Placebo compared to Baseline: Differences between mean NRS values on the last day of treatment and baseline values (BL) are shown. Dots indicate values outside of 1.5* interquartile range.

### Secondary endpoints

No statistical differences between CBDV and placebo were detectable by any of the questionnaires analysing pain characteristics, sleep quality, subjective impression of change or quality of life (**Table 2** and **Figure 4**). No significant changes in specific pain parameters in the painDETECT questionnaire were detectable. Overall, the Medication Quantification Scale (MQS) values were not significantly different between CBDV and placebo (median treatment effect of CBDV compared to placebo = 0, p=0.52, 95% CI -0.05 to 2.85; non-parametric rank sum test) (**Figure 5**). After CBDV treatment, the differences in MQS-values between baseline and end of treatment were +1.13 (SD=7.13) in the C-P groupand -0.16 (SD=0.61) in the P-C group. After placebo treatment, these differences were +0.11 (SD=3.79) and -1.87 (SD=5.26) in the C-P group and P-C group, respectively.

**Table 2.** Effect of CBDV assessed by questionnaires. painDETECT and DN4i: higher values indicate presence of neuropathic pain; Patient global impression of change (PGIC): higher values indicate a subjective improvement; all others: lower values indicate lower impairment. BPI: Brief Pain Inventory; HADS: Hospital Anxiety and Depression Scale; ISI: Insomnia Severity Index.

| Questionnaire (score range) | Effect CBDV vs. Placebo |
| --- | --- |
| **painDETECT** (0 to 38) | **-0.84** (p=0.53, 95%CI -3.59 to 1.91) |
| **DN4i** (0 to 7) | **-0.50** (p=0.18, 95%CI -1 to 0.50) |
| **BPI (pain intensity)** (0 to 10) | **+0.23** (p=0.76, 95%CI -0.63 to 1.25) |
| **BPI (influence on daily living)** (0 to 10) | **-0.35** (p=0.22, 95%CI -1.36 to 0.43) |
| **HADS (anxiety)** (0 to 21) | **-0.60** (p=0.51, 95%CI -2.44 to 1.24) |
| **HADS (depression)** (0 to 21) | **0** (p=0.91, 95%CI -1.50 to 1.50) |
| **ISI** (0 to 28) | **-1.50** (p=0.24, 95%CI -5.50 to 1) |
| **PGIC** (0 to 7) | **-0.50** (p=0.26, 95%CI -1.50 to 0.50) |

**Figure 4.**
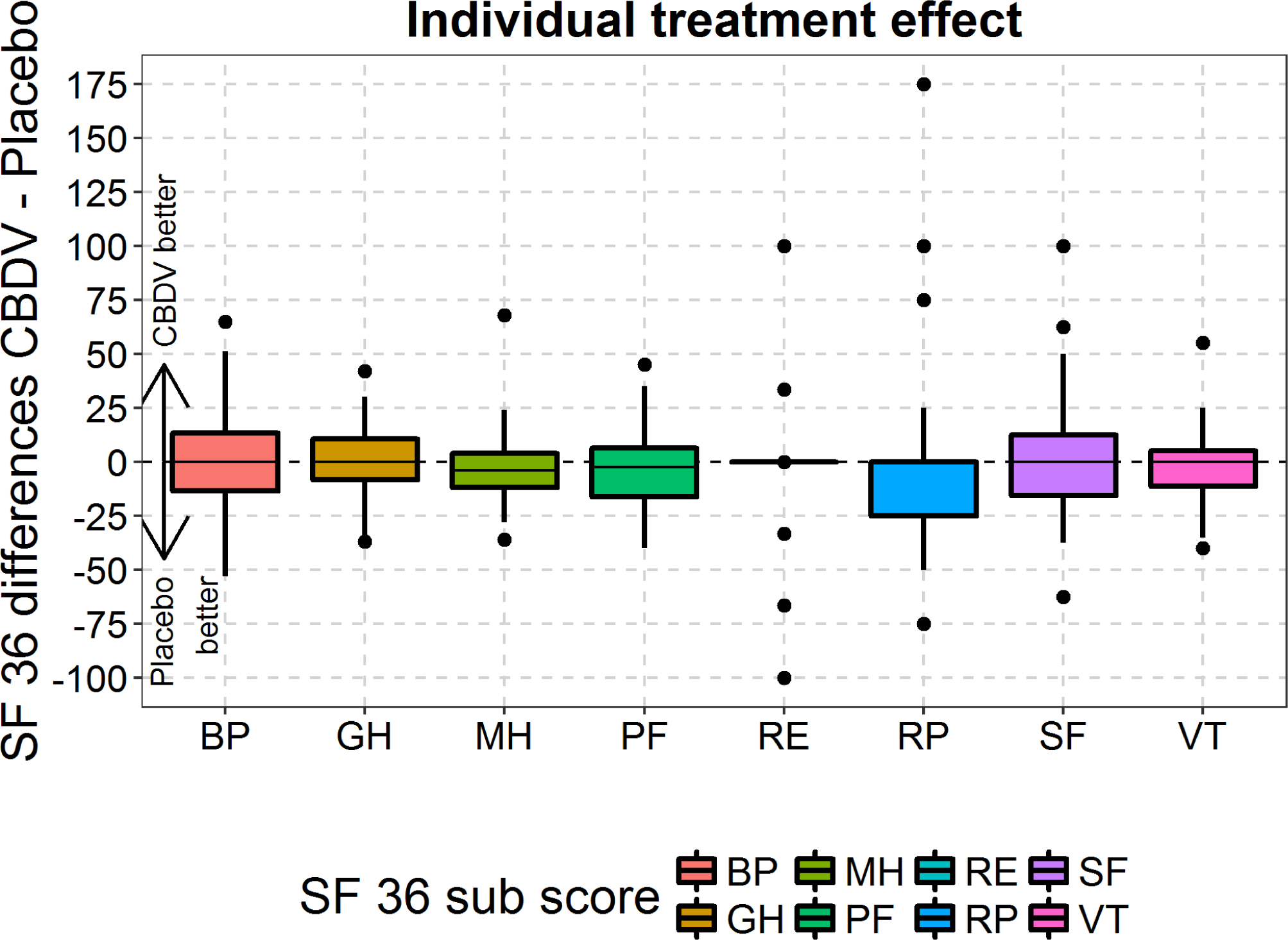
Effects by treatment sequence group – SF-36. Boxplots represent mean values; bars indicate minimum and maximum; dots indicate values outside of 1.5* interquartile range. No statistical significances were observed. Abbreviations: SF-36: 36-Item Short Form Survey; BP: Bodily Pain; GH: General Health; MH: Mental Health; PF: Physical Functioning; RE: Role Emotional; RP: Role Physical; SF: Social Functioning; VT: Vitality.

**Figure 5.**
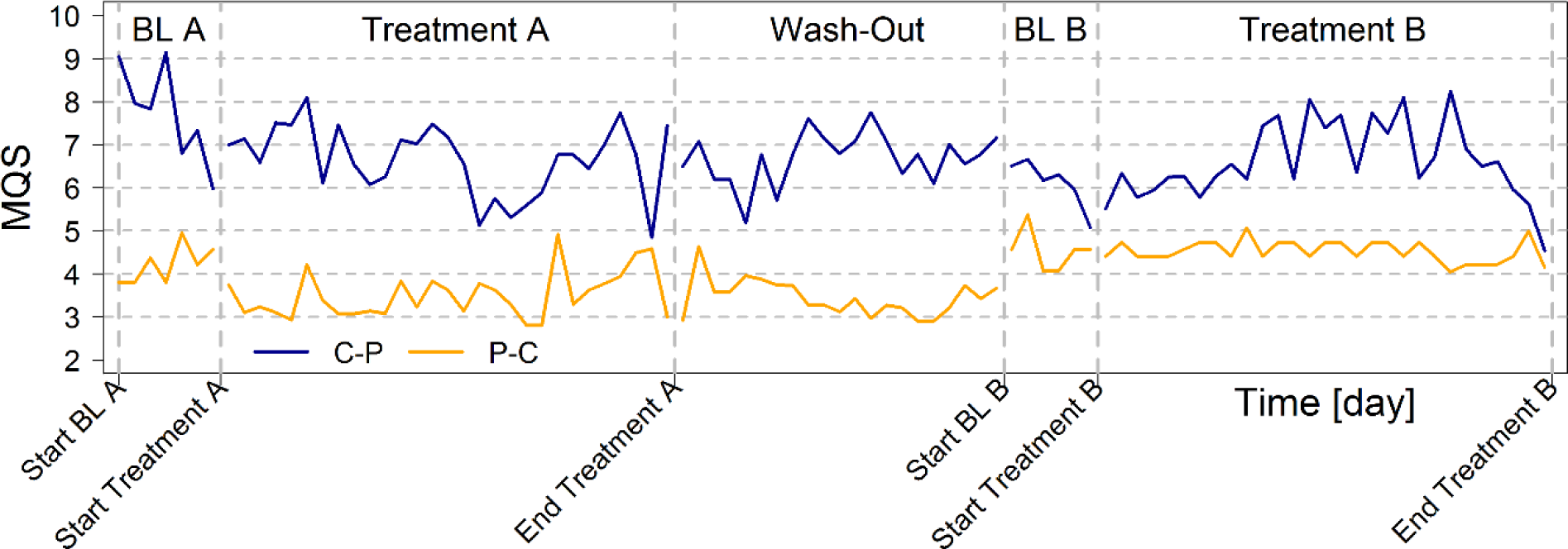
MQS-values over time: Mean MQS-values per day for each treatment sequence group. No significant differences between treatments were detected using the non-parametric rank sum test. Abbreviations: C-P = CBDV-placebo; P-C = placebo-CBDV; BL = Baseline)

### Adverse events

31 patients (91.2%) experienced at least one adverse event (AE) during CBDV treatment, whereas 27 patients (79.4%) had at least one AE during placebo. During each treatment (CBDV or placebo), 9 patients (26.5 %) experienced an AE that was considered to be related to study medication (**Table S 2**). One serious AE (acute myocardial infarction) was recorded during CBDV treatment but was judged as not related to study medication. The most common AEs were diarrhea and dry mouth (3 cases during each treatment). The incidence of AEs was similar in both treatment phases. All AEs were of low or moderate severity and only one patient withdrew study participation due to an AE (cough) during CBDV. Changes of laboratory values were considered to be not related to study medication and were not clinically relevant.

### Genetic analysis

Samples from the 28 patients who completed the entire study were genotyped using the Infinium Global Screening array (GSA24, Illumina), and whole genome sequencing was performed on this subset of patients. The small sample size did not allow a meaningful genome-wide association analysis of response, but these data will have utility in future meta-analysis efforts, and can be queried for the role of individual markers identified in other studies.

## Discussion

In this study, CBDV failed to reduce neuropathic pain intensity in HIV-patients. Additionally, we could not observe any statistically or clinically significant effects on use of supplementary pain medication, specific pain characteristics or quality of life. CBDV and placebo produced similar rates of adverse events which were of mild to moderate severity.

According to data on CB receptor knock-out mice and pharmacological studies, the mechanisms underlying analgesic effects of cannabinoids are thought to be based on the activation of CB1 and/or CB2 receptors, leading to an inhibition of pain signal transmission and/or anti-inflammatory effects (14,18,19). This may either be achieved by exogenous cannabinoids or by inhibiting enzymes degrading endocannabinoids (fatty acid amide hydrolase and monoacylglycerol lipase). Costa et al. also showed that antinociception can be produced by a cannabinoid re-uptake inhibitor in rats (20). In addition, effects of phytocannabinoids not primarily activating CB receptors have been described (21).

CBDV is mainly known for its anticonvulsant effects (22,23). Limited unpublished preclinical data indicated the occurrence of antinociceptive effects without binding to CB receptors (24). Antinociceptive effects of cannabinoids not activating CB receptors were observed in previous animal studies (25–28) but not in humans so far. Different mechanisms of action were hypothesized, such as inhibition of Diacylglycerol Lipase-α (29), another enzyme influencing endocannabinoids. Some groups observed an activation of transient receptor potentials (TRP) (29,30) and postulated that this activation could lead to desensitization of sensory neurons (31).

To evaluate the clinical effectiveness of treatments, we assessed both pain intensity and the amount of supplemental pain medication. A dose reduction of additional pain medication can minimize detrimental side effects and can therefore be useful. CBDV however, did not significantly change pain intensity or the use of additional pain medication as compared to placebo. We also examined whether CBDV can influence pain characteristics such as burning sensation, numbness or heat hyperalgesia. Due to the possible involvement of TRPV1 (29,30), a receptor which is responsible for heat sensation (37), one might assume that CBDV can alleviate burning sensations in neuropathic pain patients. In the painDETECT questionnaire however, CBDV did not influence any specific pain characteristics. To our knowledge, this is the first study investigating the influence of CBDV on such parameters.

Overall, CBDV was ineffective in our trial. The most notable (but nonsignificant) pain relief was observed in patients receiving placebo during the first phase (P-C). Since patients were randomized, this was due to chance. It is conceivable that patients who were not treated sufficiently for pain before entering our study benefitted from both treatments due to the enhanced attention in the setting of a clinical trial.

Chronic pain negatively influences many other facets of the patient’s life according to the biopsychosocial model of pain (3,32–35). Cannabinoids are known to influence emotional processes. For example, the CB receptor agonist Δ9-tetrahydrocannabinol (THC) may reduce the unpleasantness but not the intensity of pain (36). However, CBDV failed to improve any of these features in the current study.

CBDV does not bind to CB-receptors (38) and therefore should not show typical CB receptor-mediated (27) psychotropic side effects such as euphoria, reduced anxiety or feeling ’high’ (40), consistent with our findings. Since the most common side effects (diarrhea and dry mouth) did not differ between CBDV and placebo, we do not consider these AEs related to CBDV treatment. However, they could be associated with the sesame oil solution. We only observed side effects of low to moderate severity and only one patient withdrew due to such effects. For a more reliable analysis, a larger number of patients is needed.

Although we were able to obtain blood samples from most patients, this sample size was not sufficient for a meaningful genome-wide association study regarding treatment responses. However, these data are available upon request and will have utility in future meta-analysis efforts.

Due to recruitment difficulties, we could only enroll 16 patients per treatment sequence group instead of a planned sample size of 21. A larger patient population would have increased the reliability of the data. Nevertheless, we would not expect a marked change in the primary outcome since the present results are far from statistical significance. Even the lower border of the 95% CI does not promise any clinical relevance.

To conclude, this study showed that CBDV did not elicit more adverse side effects than placebo but failed to alleviate neuropathic pain or any associated condition in HIV patients. We presume that activation of CB receptors is necessary for significant analgesia. This was the first study investigating CBDV for neuropathic pain and further research with larger numbers of patients is needed.

## Methods

### Study approval, funding and registration

Trial protocol, patient information and informed consent sheets were approved by the ethics committee of the State regulatory authority Berlin (“LaGeSo”) and the German Federal Institute for Drugs and Medical Devices (“BfArM”). All patients signed written informed consent. The CONSORT guidelines, Good clinical practice (GCP) principles and the Declaration of Helsinki were strictly followed. The study was funded by the European Union as part of the consortium ’NeuroPain—Neuropathic pain: biomarkers and druggable targets within the endogenous analgesia system’ and was registered in the EudraCT-register (https://www.clinicaltrialsregister.eu/) under number 2014-005344-17.

### Study design

Data were collected from 1^st^ January 2017 to 8^th^ January 2019. We conducted a randomized, placebo-controlled, double-blind cross-over phase II trial in a single-center outpatient setting. All patients received both treatments (CBDV and placebo) in two successive phases. The order of treatments (CBDV-placebo [C-P] or placebo-CBDV [P-C]) was allocated by chance (randomized). Each patient was monitored for 13 weeks. After the screening phase, baseline values on pain scales, questionnaires and medication were recorded during a one-week phase (**Figure 6**). This was followed by a 4-week treatment phase A with either placebo or CBDV, depending on the randomization. A subsequent 3-week washout phase was included to eliminate potential carry-over effects. The duration of the wash-out phase was based on data showing an accumulation of cannabinoids in fatty tissue resulting in a half-life of about 5 days after long-term oral administration (38,41). Thereafter, another 1-week baseline phase ensued, followed by treatment phase B. Patients were then followed up for another 3 weeks. Throughout the study, the patients documented data in diaries.

**Figure 6.**
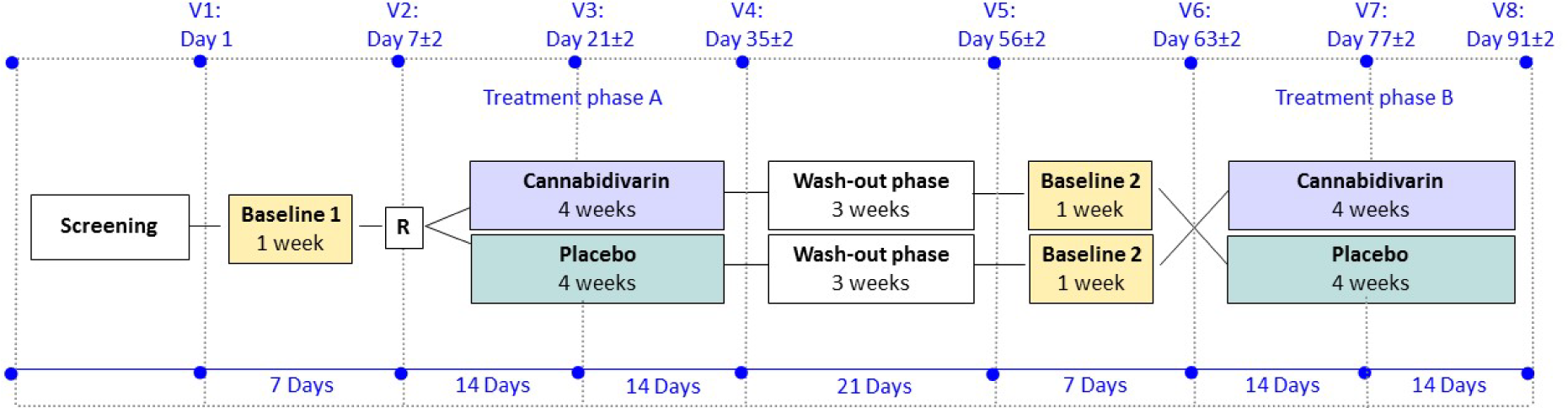
Study design. Abbreviations: V = Visit; R = Randomization

### Study participants

Participants were recruited through personal contacts to physicians and patient-advocacy groups in the greater Berlin area, as well as by advertisement in the Berlin public transportation system. For inclusion, subjects were screened for age (18-65 years), vital signs and pain intensity (≥ 4 on an 11-point numeric rating scale). The diagnosis of HIV-associated sensory neuropathy was confirmed by a clinician based on patient history, the DN4i and the Clinical HIV-associated Neuropathy Tool (CHANT). Exclusion criteria were pregnancy and lactation, major psychiatric conditions, severe diseases of the central nervous system, hepatic, renal or cardiovascular diseases or use of conventional cannabinoids, examined by blood test. Electrocardiograms were recorded at day of screening and analysed for abnormalities by an experienced cardiologist (AM). Infection with hepatitis virus B or C and AIDS-defining diseases were debarred by consulting HIV-specialists. The use of concomitant analgesics as needed was permitted throughout the study. Standard laboratory values (full blood count, liver function tests, electrolytes, glucose, urea, cholesterol, creatinine, creatinine kinase, protein and international normalized ratio) were recorded at day of screening and during the trial.

### Outcome measurements

The primary outcome was pain intensity measured thrice a day (8:30 AM, 1:00 PM and 7:00 PM) by an 11-point Numeric Rating Scale (NRS) (0 = no pain, 10 = worst pain imaginable), as documented in the patient diary. For each day, the arithmetic mean of the three NRS scores was determined. A decrease of mean NRS values by at least 20% between the last day of baseline measurement and the last day of treatment was defined as a clinically relevant effect (responder). The number of responders and non-responders to each treatment was determined. Secondary endpoints were pain characteristics, quality of life and sleep, measured by questionnaires. We used painDETECT (42), the BPI (43) and the DN4i for evaluation of pain intensity and pain characteristics (44), the HADS (45) to evaluate anxiety and depression, and the SF-36 (46), the PGIC (47) and the ISI for quality of life and sleep (48), respectively. All questionnaires were applied on the last day of each baseline phase and on the last day of each treatment phase, except PGIC, which was only used at the end of each treatment phase. Concomitant medication and side effects were recorded in the patient diary. For the analysis of concomitant pain medication we used the MQS in its 3^rd^ version, which assigns a score (on an unlimited scale) based on the detrimental effects and dose of each pain medication (49). For analysis of side effects, patients were asked to document any adverse or unusual events. These were discussed with a study physician at each visit. For standardized documentation, we used paper-based tables and classified the events with the Common Terminology Criteria for Adverse Events (CTCAE), Version 4.03.

### Randomization, allocation concealment and blinding

Randomization to the sequence of treatments occurred in blocks of four by use of paper-based, computer-generated random lists, which were stored in a locked cabinet. Patients included were pseudonymised by generating a serial number. Allocation to the treatment sequence was documented and kept in sealed envelopes. All patients and staff involved in patient contacts and assessment of outcomes were blinded until the end of the study.

## Monitoring

Two independent monitors conducted unblinded monitoring of patient safety and adherence to GCP principles throughout the trial.

### Investigational Medicinal Products (IMP)

The active agent and placebo, both dissolved in sesame oil, were identically appearing and tasting solutions. The IMP was packaged in amber-glass bottles by GW Pharmaceuticals. All bottles were subject-specific and marked with the patient-ID. The bottles with active agent contained 50 mg CBDV/ml. Patients were instructed to use 8 ml of the solution orally every morning at 9 AM, corresponding to 400 mg CBDV in the verum treatment phase (***Table 3***). The dose was chosen based on non-published preclinical and clinical phase-1-studies, carried out by GW Pharmaceuticals (38).

**Table 3.** Investigational medicinal products. Abbreviations: q.s. = quantum satis.

| Material | CBDV Solution<br>(50mg/mL) | Placebo Solution |
| --- | --- | --- |
| CBDV | 50.0 mg | - |
| Anhydrous Ethanol | 79.0mg | 79.0mg |
| Sucralose | 0.5mg | 0.5mg |
| Strawberry Flavor | 0.2mg | 0.2mg |
| Refined Sesame oil | q.s. to 1.0mL | q.s. to 1.0mL |

### Inactivation of HIV in blood samples, DNA isolation and Genetic analysis

Blood samples were obtained during the last visit from the 28 patients who gave consent for genetic analysis. 5 ml of peripheral venous blood was mixed with 15 ml of Red Cell Lysis Solution (Epicentre R) and incubated at room temperature for 10 minutes. After centrifugation, supernatant was discarded, and the pellet was dissolved in 7.5 ml Tissue and Cell Lysis Solution (Epicentre R). The solution was kept at 65°C for one hour for inactivation of HIV and cell lysis. Samples were then stored and transported at -20°C until genotyping by deCODE Genetics (Reykjavik, Iceland). Whole genome sequencing was performed by the Infinium Global Screening array (GSA24, Illumina).

### Statistical analysis

Sample size was calculated by nQuery Advisor® 7.0 based on the primary endpoint (NRS scale) and the cross-over study-design. According to previous literature, a pain reduction by 20% upon verum compared to placebo and a common standard deviation for the period differences of 2.5 seemed to be achievable and would have been clinical meaningful (50–52). We calculated that 21 patients per sequence group were sufficient to show this effect (e.g. a reduction of 20% from 6 points to 4.8 points) with a power of 85% and a two-sided type-I-error of 0.05 using a paired t-test for 2×2 crossover designs. To account for an estimated 15% dropouts, we aimed at a total of 50 patients.

Statistical analysis was based on the intent-to-treat principle. Every patient who started treatment and had at least one post-baseline measurement of the primary endpoint was included in the efficacy analysis. Continuous variables are shown as mean, standard deviation and range, while categorical parameters are given as absolute and relative frequency. For the continuous endpoints, first, the difference between sequence-specific baseline and the value after treatment was calculated. Then, for each individual the difference between the two treatment effects (CBDV - placebo) was determined. A paired t-test taking period effects into account was used for comparing the two treatments. In case of non-normality of data distribution, a non-parametric version was applied instead. 95%-confidence intervals (CI) were calculated for the treatment effects. All p values resulting from the analyses have to be considered as non-confirmatory using a cutoff of 0.05. All analyses were done using R (version 3.5.0) (53).

## Data Availability

Raw data are available at the Koordinationszentrum für Klinische Studien, Charité Universitätsmedizin Berlin, Germany

## Author contributions

SS and CS designed the study. LE, SS, MC, ML, and CS were responsible for the conduct of the trial. ÖC prepared samples for genetic studies. AM evaluated electrocardiograms. RR, LE and CS conducted data analyses. LE, RR and CS wrote the manuscript.

## Acknowledgments

The genetic analyses were carried out by Gyða Björnsdóttir and Þorgeir Þorgeirsson (deCODE Genetics, Reykjavik, Iceland). CBDV was provided by GW Pharmaceuticals (Cambridge, UK) who was not involved in trial design or data analysis. We are grateful for the continuous support by Colin Stott and Karen Twigden (GW Pharmaceuticals) and for monitoring by Dr. Susen Burock and Izabella Rauer (Charité Comprehensive Cancer Center). The study was funded by the European Commission (EU FP7-HEALTH-2013-INNOVATION-1; No. 602891-2).

## Supplementary data

**Table S 1.** Primary endpoint - NRS values. Abbreviations: C-P = CBDV-placebo; P-C = placebo-CBDV; SD = Standard Deviation; IQR = Interquartile Range.)

| Characteristic | Level | C-P | P-C | Total |
| --- | --- | --- | --- | --- |
| NRS last day base line phase 1 | n | 16 | 16 | 32 |
|  | Mean (SD) | 3.62 (1.62) | 5.23 (2.43) | 4.43 (2.19) |
|  | Median (IQR) | 3.67 (2.3; 5) | 4.83 (3.4; 6.8) | 3.67 (2.7; 6) |
|  | Range | 1.3; 6.7 | 2; 9.3 | 1.3; 9.3 |
| NRS last day treatment phase 1 | Mean (SD) | 3.29 (1.74) | 3.25 (2.4) | 3.27 (2.07) |
|  | Median (IQR) | 2.67 (2; 4.8) | 2.83 (2; 3.8) | 2.67 (2; 4.7) |
|  | Range | 0.7; 6.3 | 0; 10 | 0; 10 |
| NRS difference treatment phase 1 | Mean (SD) | -0.44 (0.57) | -1.98 (2.03) | -1.24 (1.68) |
|  | Median (IQR) | -0.33 (-0.6; -0.2) | -1.5 (-2.8; -0.9) | -0.67 (-1.7; -0.3) |
|  | Range | -1.7; 0.3 | -6; 1.3 | -6; 1.3 |
| Pain reduction ≥ 20% treatment phase 1 | | 4 | 13 | 17 |
| NRS last day baseline phase 2 | Mean (SD) | 3.3 (1.98) | 3.88 (2.69) | 3.59 (2.34) |
|  | Median (IQR) | 2.83 (1.8; 4.4) | 3.33 (2.3; 5.2) | 3.17 (2.2; 4.8) |
|  | Range | 0.3; 7 | 0.3; 10 | 0.3; 10 |
| NRS last day treatment phase 2 | Mean (SD) | 3.01 (2.14) | 3.57 (1.92) | 3.29 (2.02) |
|  | Median (IQR) | 2.33 (2; 3.2) | 3.5 (2.5; 4.8) | 3 (2; 4.2) |
|  | Range | 0; 8 | 0.7; 7.3 | 0; 8 |
| NRS difference treatment phase 2 | Mean (SD) | -0.29 (0.99) | -0.31 (1.82) | -0.3 (1.44) |
|  | Median (IQR) | -0.33 (-1.2; 0.5) | -0.33 (-0.9; 0.2) | -0.33 (-1.1; 0.3) |
|  | Range | -1.5; 1.7 | -3.3; 4.7 | -3.3; 4.7 |
| Pain reduction ≥ 20% treatment phase 2 | | 6 | 5 | 11 |

**Table S 2.** Adverse events related to study medication.

| AE | Number of patients |  |
| --- | --- | --- |
|  | CBDV | Placebo |
| Concentration disorders | 1 | 1 |
| Constipation | 0 | 1 |
| Diarrhea | 3 | 1 |
| Dizziness | 0 | 2 |
| Dry mouth | 3 | 5 |
| Dysesthesia | 0 | 1 |
| Dyspepsia | 0 | 1 |
| Fatigue | 0 | 1 |
| Gastrointestinal disorders | 0 | 1 |
| Headache | 1 | 3 |
| Hyperhidrosis | 0 | 2 |
| Hypertrichosis | 1 | 0 |
| Insomnia | 0 | 1 |
| Mood disturbances | 0 | 1 |
| Nausea | 0 | 2 |
| Numbness in neck | 1 | 0 |
| Pruritus | 1 | 1 |
| Tachycardia | 0 | 1 |
| Vision disorders | 0 | 1 |

**Figure S 1.**
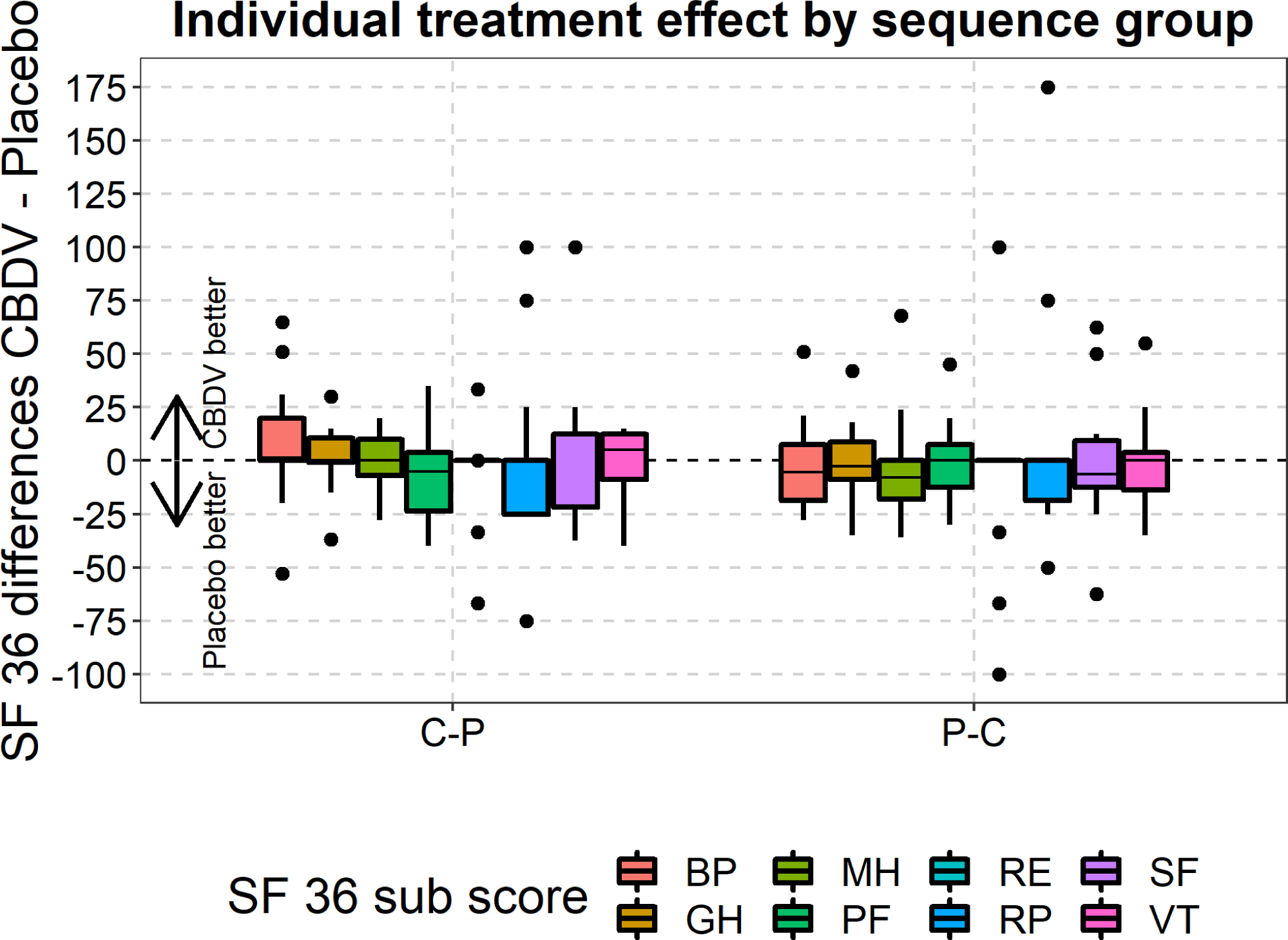
Quality of life assessed by SF-36, differentiated by treatment groups. Abbreviations are explained in Figure 4. Boxplots represent mean values; bars indicate minimum and maximum; dots indicate values outside of 1.5* interquartile range.

